# Serotype distribution among adults with community-acquired pneumococcal pneumonia in Japan between 2019 and 2022: A multicenter observational study

**DOI:** 10.1101/2025.01.29.25321300

**Authors:** Haruka Maeda, Isao Ito, Eiichiro Sando, Nobuyoshi Hamao, Masahiro Shirata, Bhim Gopal Dhoubhadel, Desmond Opoku Ntiamoah, Issei Oi, Kensuke Nishioka, Hiroshi Fujii, Kayoko Okamura, Taisei Inoue, Takashi Yamada, Seisuke Niibayashi, Mitsuhiro Tsukino, Yuya Fujii, Michiko Tsuchiya, Yasuharu Nakahara, Yoshinori Hasegawa, Atsushi Nakagawa, Takakazu Sugita, Akihiro Ito, Naoki Sakai, Yusuke Kaji, Yuko Toyoda, Tomoyuki Urata, Norichika Asoh, Akira Nishiyama, Ai Yagiuchi, Toru Morikawa, Atsuhito Ushiki, Masayuki Ishida, Konosuke Morimoto

**Author notes:** First Department of Internal Medicine, Shinshu University School of Medicine, Japan (Atsuhito Ushiki). Department of Respiratory Medicine, Chikamori Hospital, Japan (Masayuki Ishida).

## Abstract

**Background:** *Streptococcus pneumoniae* is a leading cause of community-acquired pneumonia in adults. With the introduction of pneumococcal conjugate vaccines (PCVs) into pediatric national immunization programs, the serotype distribution of pneumococcal disease among adults has changed due to herd immunity. In Japan, PCV15 and PCV20 have been introduced, and PCV21 has been under review for approval in adults. This study aimed to assess the distribution of pneumococcal serotypes among adults with pneumococcal pneumonia in Japan between May 2019 and December 2022.

**Methods:** This multicenter observational study enrolled patients aged ≥18 years with community-acquired, culture-positive pneumococcal pneumonia from May 2019 to December 2022. Pneumococcal isolates were serotyped using the Quellung reaction, and proportions of individual and vaccine-covered serotypes were analyzed.

**Results:** A total of 583 adult patients with pneumococcal pneumonia were included. The median age was 74 years (interquartile range: 66–82 years), 383 (65.7%) patients were male, and 387 (66.4%) patients had one or more underlying medical conditions. The most common serotypes were serotype 3 (12.5%), 35B (12.0%), 15A (7.7%), 11A (6.7%), and 23A (6.3%). The proportion of serotypes covered by PCV13, PCV15, PCV20, PPSV23, and PCV21 were 24.0, 28.0, 43.7, 44.1, and 71.9%, respectively. The proportions of vaccine-covered serotypes were similar between patients aged <65 and ≥65 years. Notably, serotype 3 was more prevalent among patients living in nursing homes (25.9%) compared with those living at home (11.2%).

**Conclusion:** Serotypes 3 and 35B were the most common in adults with pneumococcal pneumonia in Japan during the study period. The proportions of PCV20- and PCV21-covered serotypes suggest that these new vaccines may offer additional protection against adult pneumococcal pneumonia. With the availability of newly developed PCVs for adults in Japan, it is time to reassess the optimal pneumococcal vaccination policy for adults.

## 1. Introduction

Community-acquired pneumonia (CAP) is a leading cause of morbidity and mortality worldwide, particularly among children under five years of age and older adults (1, 2). *Streptococcus pneumoniae* is a major causative pathogen of CAP. Although *S. pneumoniae* can cause invasive pneumococcal disease (IPD), which progresses to more severe diseases, the burden of pneumococcal CAP among older adults is marked, as the incidence of pneumococcal pneumonia is higher than that of IPD (3, 4). Therefore, when discussing pneumococcal vaccination policies, it is essential to consider epidemiological evidence for both IPD and pneumococcal pneumonia. The introduction of pneumococcal conjugate vaccines (PCVs) into pediatric national immunization programs (NIP) has significantly reduced pneumococcal disease caused by vaccine-covered serotypes in both adults and children who have not received vaccines due to herd immunity (5–7). PCVs have effectively reduced pneumococcal nasopharyngeal carriage, thereby limiting *S. pneumoniae* transmission from children to adults. However, pneumococcal disease caused by serotype 3 have persisted, and serotypes not covered by PCVs have increased in prevalence (8, 9).

Recently, two new higher-valency PCVs, the 15-valent pneumococcal conjugate vaccine (PCV15) and 20-valent pneumococcal conjugate vaccine (PCV20), were approved and introduced in Japan (10, 11). These vaccines have replaced the 13-valent pneumococcal conjugate vaccine (PCV13) in the pediatric NIP (10). Both PCV15 and PCV20 are also available for adults; however, they are not included in NIP for older adults. In addition, the 21-valent pneumococcal conjugate vaccine (PCV21), which targets residual serotypes affecting adults, is currently under review for approval in Japan (12). With multiple pneumococcal vaccine options now available for adults, the pneumococcal vaccination policy in Japan requires reconsideration. However, data on the serotype distribution of pneumococcal pneumonia in adults in Japan are limited.

This study aimed to assess the distribution of individual pneumococcal serotypes and proportions of vaccine-covered serotypes among adults with pneumococcal pneumonia in Japan between 2019 and 2022.

## 2. Methods

### 2.1. Pneumococcal vaccination policy and coverage in Japan

The Japanese government introduced health insurance-based coverage for the 23-valent pneumococcal polysaccharide vaccine (PPSV23) in 1992 for asplenic or splenectomized individuals aged ≥2 years. In 2014, PPSV23 was incorporated into NIP for older adults aged ≥65 years (13). PCV13 became available for adults aged ≥65 years in 2014 (13), followed by PCV15 in 2022 and PCV20 in 2024. However, none of these PCVs have been included in NIP to date (12).

Currently, for adults who have not received PPSV23, the Joint Committee of the Japanese Respiratory Society, Japanese Association for Infectious Diseases, and Japanese Society for Vaccinology recommend PPSV23 as routine vaccination for those aged 65 years. For those aged >65 years, recommended optional vaccinations are: PPSV23, PCV20, or a sequential strategy involving PCV15 followed by PPSV23. For adults who have already received PPSV23, the recommendations are PPSV23, PCV20, or the sequential strategy involving PCV15 followed by PPSV23 (Supplementary Figure 1) (11).

For pediatric vaccination, the 7-valent pneumococcal conjugate vaccine (PCV7) was introduced into NIP in April 2013 and was replaced by PCV13 in July 2013. PCV15 was incorporated into the pediatric NIP in April 2024, followed by PCV20 in October 2024. Currently, PCV13 is no longer used in the pediatric NIP (10, 14). The routine pediatric vaccination schedule for PCVs consists of a three-dose primary series and booster dose, administered between two months and five years of age. From 2018 to 2022, vaccination coverage for children receiving PCVs, including the booster dose, was high at 94–100% (15). In contrast, vaccination coverage with PPSV23 for adults aged ≥65 years was low, at less than 30% (15, 16).

### 2.2. Study design and participants

The Japan Pneumococcal Vaccine Effectiveness 2 (JPAVE2) study is a multicenter observational study designed to evaluate the pneumococcal serotype distribution in adults with pneumococcal pneumonia. This study was conducted through collaboration between Nagasaki University, Kyoto University, and participating hospitals: 24 hospitals in 13 prefectures across Japan’s three main islands (Supplementary Figure 2). The analysis included patients enrolled between May 2019 and December 2022, encompassing an 11-month period before the start of the coronavirus disease 2019 (COVID-19) pandemic in Japan.

Adults with community-acquired, culture-positive pneumococcal pneumonia were eligible for enrollment. Eligibility criteria were as follows: age ≥18 years; clinical signs and symptoms consistent with pneumonia, such as fever, chills or rigors, cough, sputum, pleuritic chest pain, dyspnea, or tachypnea; new pulmonary infiltrates detected on chest X-ray or computed tomography; and isolation of *S. pneumonia* from sputum, blood, or pleural effusion cultures obtained during routine clinical practice. Both hospitalized patients and outpatients were included in the study. Patients who developed pneumonia more than 48 hours after hospital admission were excluded.

### 2.3. Data collection

Demographic and clinical data were collected from medical records using a standardized data collection form. Pneumococcal vaccination history was obtained from patients, their family members, or medical records. Pneumonia severity at the time of the initial visit or hospital admission was assessed using the CURB-65 score (17, 18). When calculating this score, SpO2 <90% or the need for oxygen administration was used instead of tachypnea, as respiratory failure is often assessed based on SpO2 rather than respiratory rate in Japan (19). Moderate and severe pneumonia were defined as CURB-65 scores of 2 and ≥3, respectively.

### 2.4. Pneumococcal serotype categories

Pneumococcal serotypes were grouped as follows: PCV13 included serotype 4, 6 B, 9V, 14, 18C, 19F, 23F, 1, 5, 7F, 3, 6A, and 19A; PCV15 included PCV13 plus serotype 22F and 33F; PCV20 included PCV15 plus serotype 8, 10A, 11A, 12F, and 15 B; PPSV23 included serotype 1, 2, 3, 4, 5, 6 B, 7F, 8, 9N, 9V, 10A, 11A, 12F, 14, 15 B, 17F, 18C, 19A, 19F, 20, 22F, 23F, and 33F. PCV21 included serotype 3, 6A, 7F, 19A, 22F, 33F, 8, 10A, 11A, 12F, 9N,

17F, 20, 15A, 15C, 16F, 23A, 23B, 24F, 31, and 35B (Supplementary Figure 3).

### 2.5. Microbiological testing

*S. pneumoniae* isolates were stored in a cryotube containing 1.0 mL of STGG medium (skim milk: 2 g, tryptone: 3 g, glucose: 0.5 g, glycerol: 10 mL, distilled water: 100 mL) at -80℃ (20). The isolates were then transported from each participating hospital to the Institute of Tropical Medicine, Nagasaki University. At Nagasaki University, optochin-sensitivity and bile solubility tests were performed to identify *S. pneumoniae*, and serotyping was conducted using the Quellung reaction (ImmuLex Pneumotest, SSI Diagnostica, Hillerod, Denmark) (21).

### 2.6. Statistical analysis

Baseline characteristics and clinical information are summarized. Continuous variables are presented as the median and interquartile range (IQR), and categorical variables are expressed as numbers and proportions. The proportions of individual pneumococcal and vaccine-covered serotypes were analyzed overall as well as separately for patients aged <65 and ≥65 years, for the enrollment period both before (May 2019 to March 2020) and during (April 2020 to December 2022) the COVID-19 pandemic. In addition, we reported the proportions of individual and vaccine-covered serotypes of patients living in nursing homes and those living at home. To compare the proportions of individual or vaccine-covered serotypes between the two groups, such as by age group (<65 years or ≥65 years) and enrollment period (before or during the COVID-19 pandemic), the chi-square test was employed. A two-sided p-value of <0.05 was considered significant. All analyses were performed using Stata version 16 (Stata Corp., College Station, TX, USA).

### 2.7. Ethics

This study was approved by the Institutional Review Board of the Institute of Tropical Medicine, Nagasaki University (approval no.: 180608191), and the participating hospitals.

## 3. Results

### 3.1. Demographics and clinical characteristics

Between May 2019 and December 2022, a total of 583 adult patients with pneumococcal pneumonia were included in the study. *S. pneumoniae* was isolated from sputum, blood, and pleural effusion in 94.5% (551/583), 8.1% (47/583), and 0.2% (1/583) of the patients, respectively. In 2.7% (16/583) of the patients, *S. pneumoniae* was identified in both sputum and blood, with the same pneumococcal serotype detected. Among the total patients, 451 (77.4%) were aged ≥65 years, with the median age was 74 years (IQR: 66–82 years); 383 (65.7%) patients were male, 54 (9.3%) lived in nursing homes, and 352 (60.4%) had a history of smoking (Table 1). At least one underlying medical condition was observed in 387 (66.4%) patients. Overall, 425 patients (72.9%) were hospitalized, 305 (52.3%) had a CURB-65 score ≥2, and 221 (37.9%) presented with SpO2 <90% or required oxygen administration at the time of the hospital visit. In-hospital deaths were recorded in 31 (5.3%). Among patients aged ≥65 years with a known PPSV23 vaccination status, 77 of 264 (29.2%) had received PPSV23 within five years before the hospital visit.

**Table 1.**
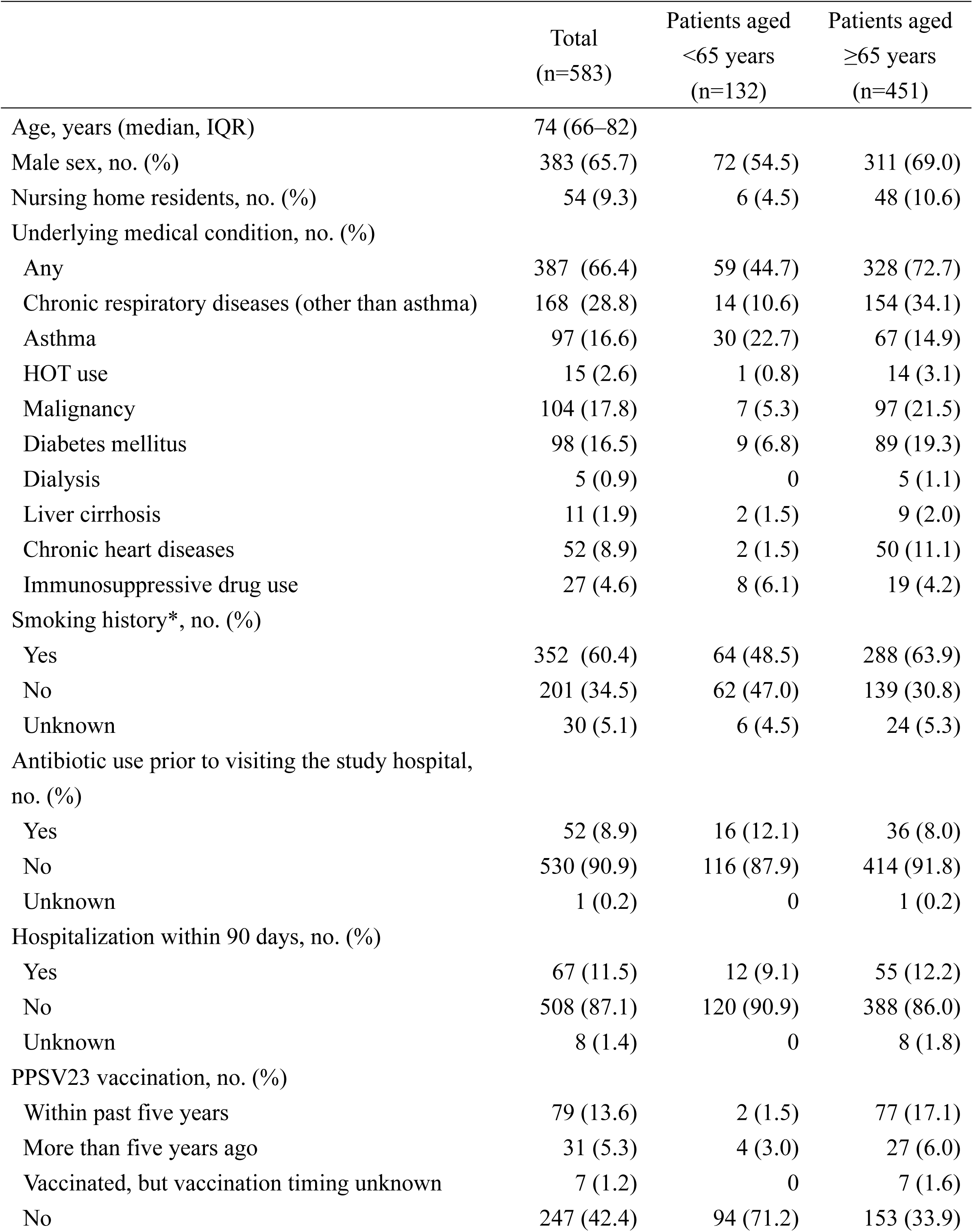
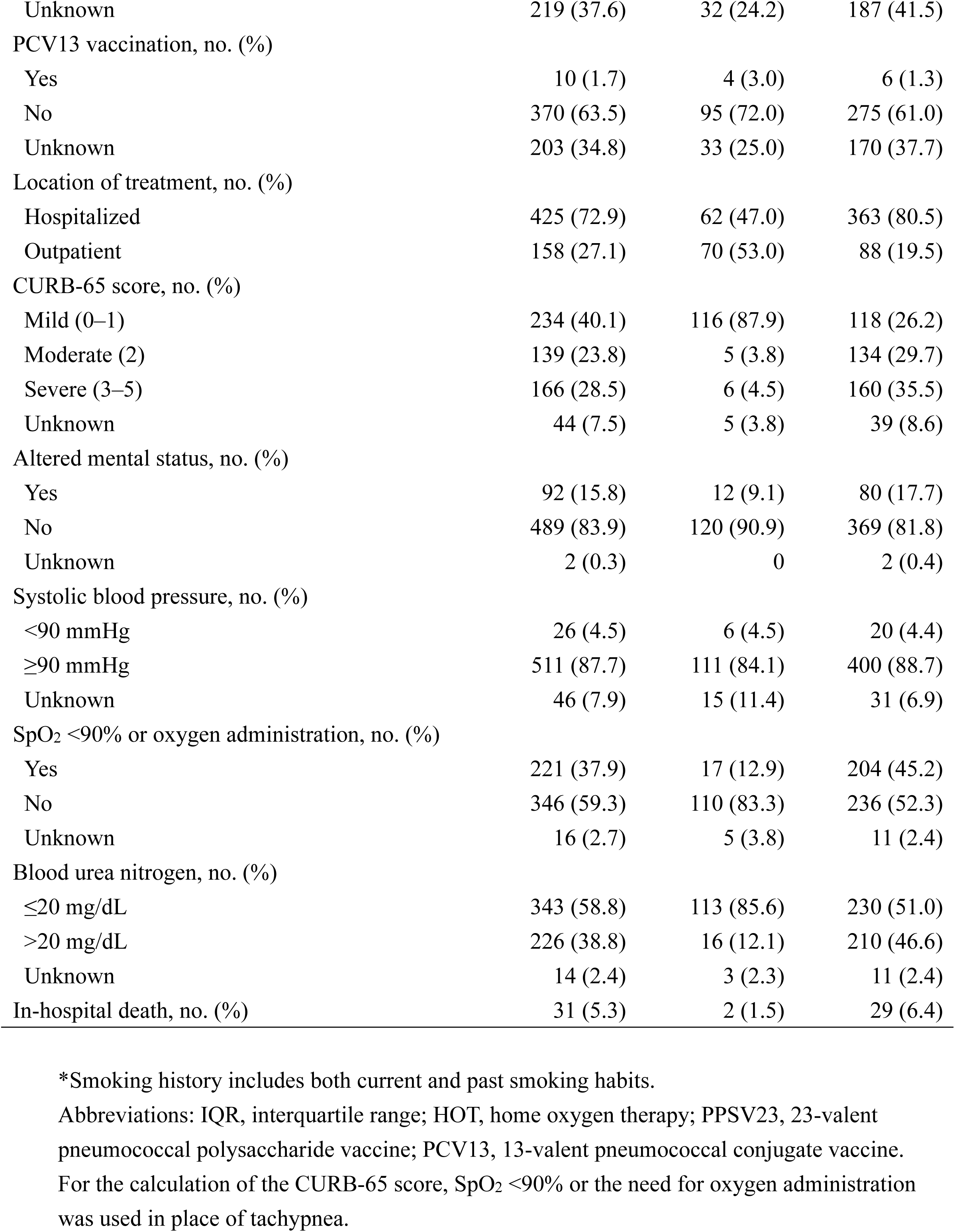
Demographics and clinical characteristics of adult pneumococcal pneumonia patients in Japan between May 2019 and December 2022.

|  | Total<br>(n=583) | Patients aged<br><65 years<br>(n=132) | Patients aged<br>≥65 years<br>(n=451) |
| --- | --- | --- | --- |
| Age, years (median, IQR) | 74 (66–82) |  |  |
| Male sex, no. (%) | 383 (65.7) | 72 (54.5) | 311 (69.0) |
| Nursing home residents, no. (%) | 54 (9.3) | 6 (4.5) | 48 (10.6) |
| Underlying medical condition, no. (%) |  |  |  |
| Any | 387 (66.4) | 59 (44.7) | 328 (72.7) |
| Chronic respiratory diseases (other than asthma) | 168 (28.8) | 14 (10.6) | 154 (34.1) |
| Asthma | 97 (16.6) | 30 (22.7) | 67 (14.9) |
| HOT use | 15 (2.6) | 1 (0.8) | 14 (3.1) |
| Malignancy | 104 (17.8) | 7 (5.3) | 97 (21.5) |
| Diabetes mellitus | 98 (16.5) | 9 (6.8) | 89 (19.3) |
| Dialysis | 5 (0.9) | 0 | 5 (1.1) |
| Liver cirrhosis | 11 (1.9) | 2 (1.5) | 9 (2.0) |
| Chronic heart diseases | 52 (8.9) | 2 (1.5) | 50 (11.1) |
| Immunosuppressive drug use | 27 (4.6) | 8 (6.1) | 19 (4.2) |
| Smoking history*, no. (%) |  |  |  |
| Yes | 352 (60.4) | 64 (48.5) | 288 (63.9) |
| No | 201 (34.5) | 62 (47.0) | 139 (30.8) |
| Unknown | 30 (5.1) | 6 (4.5) | 24 (5.3) |
| Antibiotic use prior to visiting the study hospital,<br>no. (%) |  |  |  |
| Yes | 52 (8.9) | 16 (12.1) | 36 (8.0) |
| No | 530 (90.9) | 116 (87.9) | 414 (91.8) |
| Unknown | 1 (0.2) | 0 | 1 (0.2) |
| Hospitalization within 90 days, no. (%) |  |  |  |
| Yes | 67 (11.5) | 12 (9.1) | 55 (12.2) |
| No | 508 (87.1) | 120 (90.9) | 388 (86.0) |
| Unknown | 8 (1.4) | 0 | 8 (1.8) |
| PPSV23 vaccination, no. (%) |  |  |  |
| Within past five years | 79 (13.6) | 2 (1.5) | 77 (17.1) |
| More than five years ago | 31 (5.3) | 4 (3.0) | 27 (6.0) |
| Vaccinated, but vaccination timing unknown | 7 (1.2) | 0 | 7 (1.6) |
| No | 247 (42.4) | 94 (71.2) | 153 (33.9) |
| Unknown | 219 (37.6) | 32 (24.2) | 187 (41.5) |
| PCV13 vaccination, no. (%) |  |  |  |
| Yes | 10 (1.7) | 4 (3.0) | 6 (1.3) |
| No | 370 (63.5) | 95 (72.0) | 275 (61.0) |
| Unknown | 203 (34.8) | 33 (25.0) | 170 (37.7) |
| Location of treatment, no. (%) |  |  |  |
| Hospitalized | 425 (72.9) | 62 (47.0) | 363 (80.5) |
| Outpatient | 158 (27.1) | 70 (53.0) | 88 (19.5) |
| CURB-65 score, no. (%) |  |  |  |
| Mild (0–1) | 234 (40.1) | 116 (87.9) | 118 (26.2) |
| Moderate (2) | 139 (23.8) | 5 (3.8) | 134 (29.7) |
| Severe (3–5) | 166 (28.5) | 6 (4.5) | 160 (35.5) |
| Unknown | 44 (7.5) | 5 (3.8) | 39 (8.6) |
| Altered mental status, no. (%) |  |  |  |
| Yes | 92 (15.8) | 12 (9.1) | 80 (17.7) |
| No | 489 (83.9) | 120 (90.9) | 369 (81.8) |
| Unknown | 2 (0.3) | 0 | 2 (0.4) |
| Systolic blood pressure, no. (%) |  |  |  |
| <90 mmHg | 26 (4.5) | 6 (4.5) | 20 (4.4) |
| ≥90 mmHg | 511 (87.7) | 111 (84.1) | 400 (88.7) |
| Unknown | 46 (7.9) | 15 (11.4) | 31 (6.9) |
| SpO<sub>2</sub> <90% or oxygen administration, no. (%) |  |  |  |
| Yes | 221 (37.9) | 17 (12.9) | 204 (45.2) |
| No | 346 (59.3) | 110 (83.3) | 236 (52.3) |
| Unknown | 16 (2.7) | 5 (3.8) | 11 (2.4) |
| Blood urea nitrogen, no. (%) |  |  |  |
| ≤20 mg/dL | 343 (58.8) | 113 (85.6) | 230 (51.0) |
| >20 mg/dL | 226 (38.8) | 16 (12.1) | 210 (46.6) |
| Unknown | 14 (2.4) | 3 (2.3) | 11 (2.4) |
| In-hospital death, no. (%) | 31 (5.3) | 2 (1.5) | 29 (6.4) |

\*Smoking history includes both current and past smoking habits.
Abbreviations: IQR, interquartile range; HOT, home oxygen therapy; PPSV23, 23-valent pneumococcal polysaccharide vaccine; PCV13, 13-valent pneumococcal conjugate vaccine.
For the calculation of the CURB-65 score, SpO<sub>2</sub> <90% or the need for oxygen administration was used in place of tachypnea.

### 3.2. Individual pneumococcal serotype distribution among adult pneumococcal pneumonia patients

In the 583 adult pneumococcal pneumonia patients, the most common serotype was serotype 3 (12.5%), followed by serotype 35B (12.0%), 15A (7.7%), 11A (6.7%), and 23A (6.3%) (Figure 1, Supplementary Table 1). Non-typable *S. pneumoniae* was isolated in 2.2% (13/583) patients. For patients aged <65 years, serotype 35B (12.9%) was the most common, followed by serotype 3 (12.1%) and 11A (9.1%). Among patients aged ≥65 years, serotype 3 (12.6%) was the most common, followed by serotype 35B (11.8%) and 15A (8.2%). The distribution of individual serotypes was similar in the two groups.

**Figure 1.**
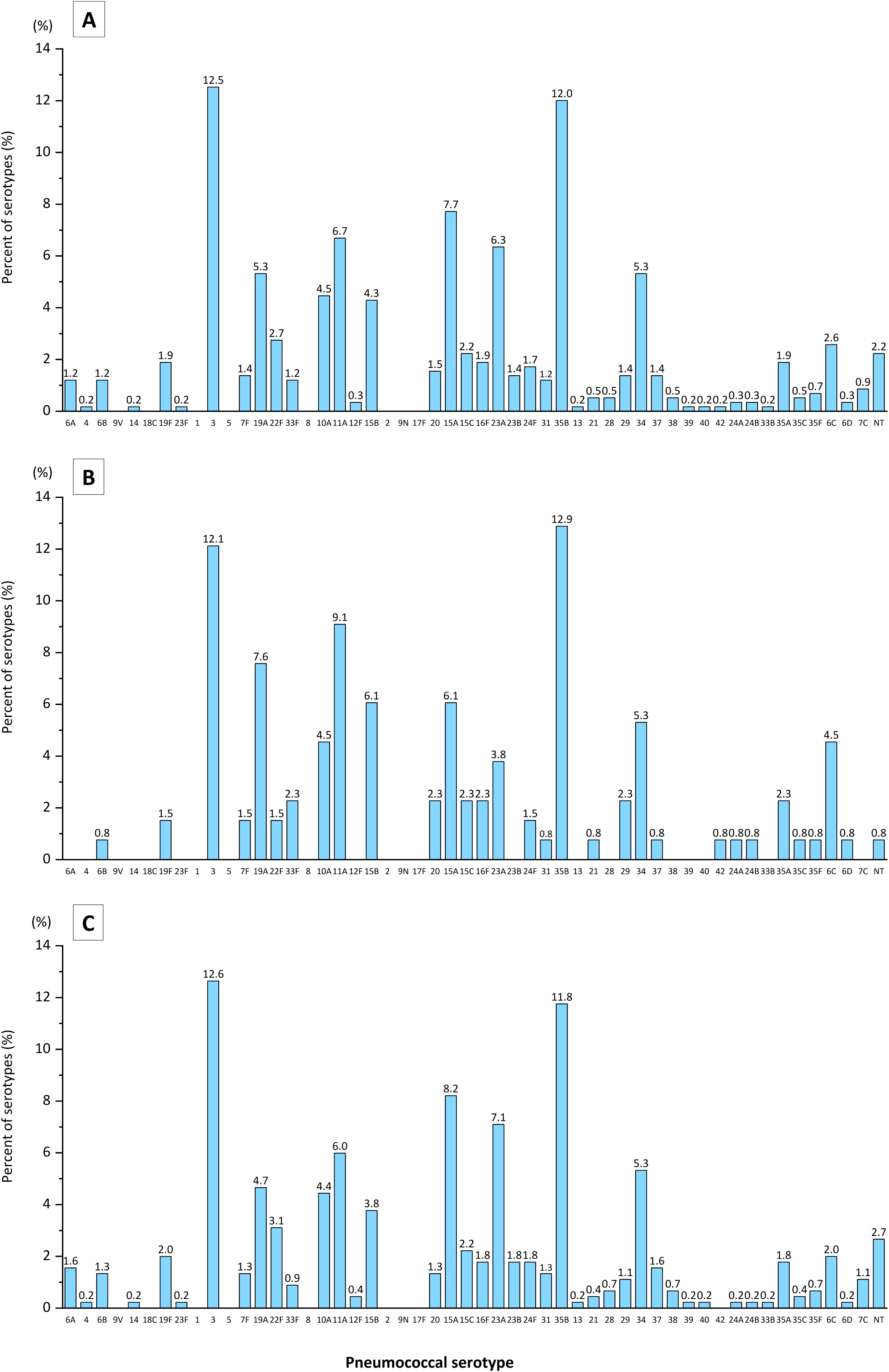
Pneumococcal serotypes detected in adult pneumococcal pneumonia patients. Bars represent the percentage of detected pneumococcal serotypes. A. Pneumococcal serotypes detected in adult patients aged ≥18 years. B. Pneumococcal serotypes detected in adult patients aged 18–64 years. C. Pneumococcal serotypes detected in adult patients aged ≥65 years.

### 3.3. Proportions of vaccine-covered serotypes among adult pneumococcal pneumonia patients

Figure 2 and Supplementary Table 2 show the proportions of vaccine-covered serotypes among adult pneumococcal pneumonia patients. PCV13 serotype was detected in 140 patients (24.0%), with serotype 3 and 19A accounting for 52.1 and 22.1% of PCV13 serotype, respectively. PCV15, PCV20, and PPSV23 serotype were detected in 163 (28.0%), 255 (43.7%), and 257 (44.1%) patients, respectively. Serotypes included in PCV21 accounted for 71.9% (419/583) of all patients, while serotypes included in PCV21 but not PCV20 represented 36.0% (210/583). Serotype 35B constituted 33.3% (70/210) of the serotypes included in PCV21 but not PCV20. The proportions of vaccine-covered serotypes were similar between patients aged <65 and ≥65 years. When comparing the proportions of vaccine-covered serotypes before and during the COVID-19 pandemic, no significant differences were observed between the two groups (Supplementary Table 3; Supplementary Figures 6 and 7).

**Figure 2.**
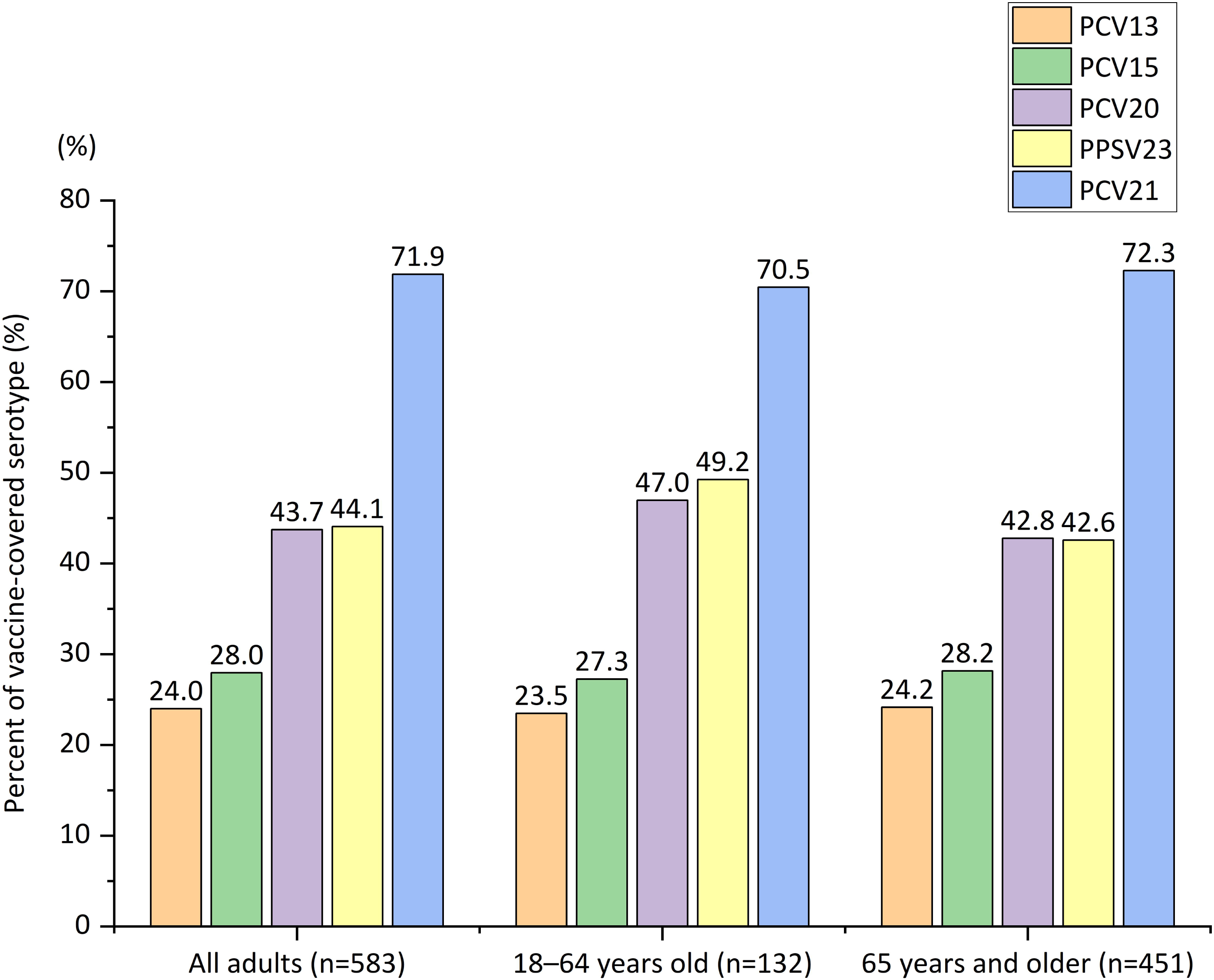
Percentage of adult pneumococcal pneumonia patients with serotypes included in PCV13, PCV15, PCV20, PPSV23, and PCV21, overall and by age group. Bars represent the percentage of pneumococcal pneumonia patients with pneumococcal serotypes included in each vaccine. PCV13 serotype include serotype 4, 6 B, 9V, 14, 18C, 19F, 23F, 1, 5, 7F, 3, 6A, and 19A; PCV15 serotype include PCV13 serotype plus serotype 22F and 33F; PCV20 serotype include PCV15 serotype plus serotype 8, 10A, 11A, 12F, and 15 B; PPSV23 serotype include serotype 1, 2, 3, 4, 5, 6 B, 7F, 8, 9N, 9V, 10A, 11A, 12F, 14, 15 B, 17F, 18C, 19A, 19F, 20, 22F, 23F, and 33F. PCV21 serotype include serotype 3, 6A, 7F, 19A, 22F, 33F, 8, 10A, 11A, 12F, 9N, 17F, 20, 15A, 15C, 16F, 23A, 23B, 24F, 31, and 35B. Abbreviations: PCV13, 13-valent pneumococcal conjugate vaccine; PCV15, 15-valent pneumococcal conjugate vaccine; PCV20, 20-valent pneumococcal conjugate vaccine; PPSV23, 23-valent pneumococcal polysaccharide vaccine; PCV21, 21-valent pneumococcal conjugate vaccine.

### 3.4. Pneumococcal serotype distribution among patients living at home and in nursing homes

Among patients living in nursing homes, serotype 3 was the most common, accounting for 25.9% (14/54), being more than two-times the proportion in patients living at home (11.2%, 59/529). In contrast, serotype 35B was the most common serotype among patients living at home (12.7%, 67/529), whereas its proportion was lower among patients living in nursing homes (5.6%, 3/54) (Figure 3, Supplementary Table 1). PCV20 and PCV21 serotypes accounted for 50.0 and 66.7% of patients living in nursing homes, respectively, and 43.1 and 72.4% of patients living at home, respectively (Supplementary Figure 4). While the proportions of PCV20 and PCV21 serotypes were comparable between the two groups, significant differences were observed in the proportions of serotype 3 and other serotypes included in both PCV20 and PCV21 (p-value <0.05) (Figure 4).

**Figure 3.**
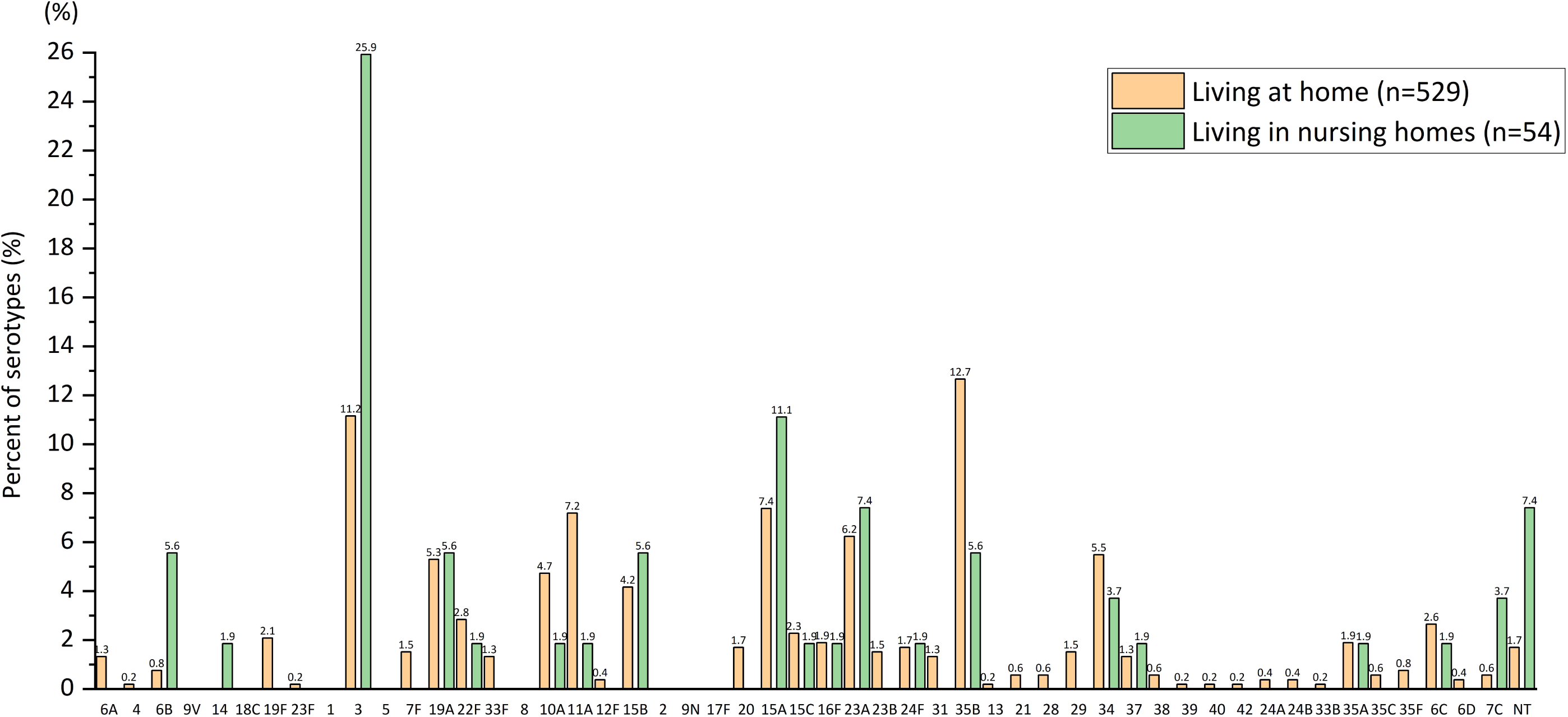
Pneumococcal serotypes detected in adult pneumococcal pneumonia patients living at home and those living in nursing homes. Orange bars represent the percentage of detected pneumococcal serotypes in patients living at home, while green bars represent that in patients living in nursing homes.

**Figure 4.**
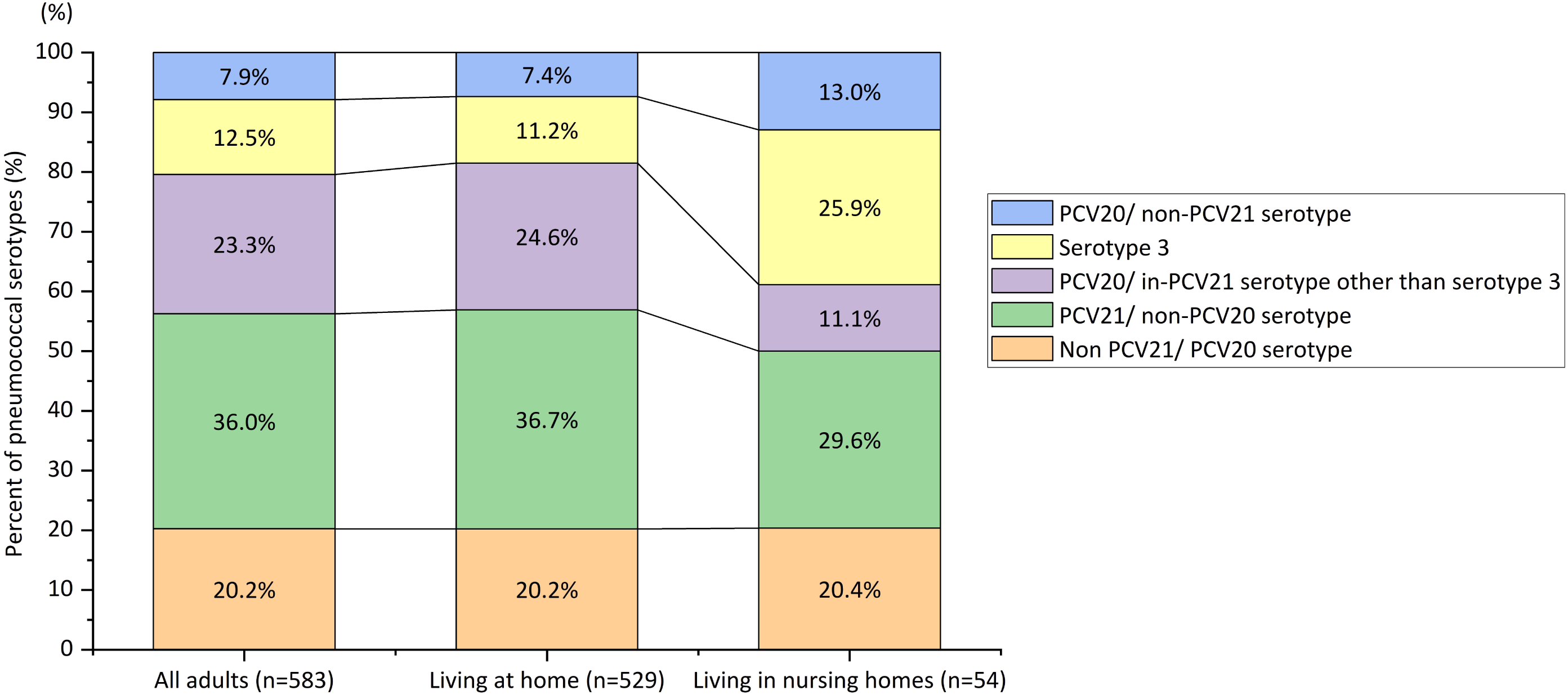
Percentage of adult pneumococcal pneumonia patients with serotype 3 and vaccine-covered serotypes, overall, in patients living at home, and in patients living in nursing homes. PCV20/non-PCV21 include serotype 1, 4, 5, 6B, 9V, 14, 18C, 19F, 23F, and 15B. PCV20/in-PCV21 other than serotype 3 include serotype 6A, 7F, 19A, 22F, 33F, 8, 10A, 11A, and 12F. PCV21/non-PCV20 include serotype 9N, 17F, 20, 15A, 15C, 16F, 23A, 23B, 24F, 31, and 35B. Non-PCV21/PCV20, which are not included in PCV20 or PCV21, include serotype 2, 13, 21, 28, 29, 34, 37, 38, 39, 40, 42, 24A, 24B, 33B, 35A, 35C, 35F, 6C, 6D, 7C, and non-typable. Abbreviations: PCV20, 20-valent pneumococcal conjugate vaccine; PCV21, 21-valent pneumococcal conjugate vaccine.

## 4. Discussion

In this study, we described the pneumococcal serotype distribution in adult pneumococcal pneumonia patients in Japan between May 2019 and December 2022. The key findings were as follows: (1) serotype 3 was the most common overall, followed by serotype 35B, 15A, 11A, and 23A; (2) PCV15, PCV20, PPSV23, and PCV21 serotypes accounted for 28.0, 43.7, 44.1, and 71.9% of patients aged ≥18 years, respectively, and 28.2, 42.8, 42.6, and 72.3% of patients aged ≥65 years, respectively; (3) the proportion of serotype 3 was notably higher among patients living in nursing homes (25.9%) compared with those living at home (11.2%), even though the proportions of PCV20 and PCV21 serotypes were similar in the two groups; (4) there was no significant difference in the proportions of vaccine-covered serotypes between before and during the COVID-19 pandemic.

With the introduction of PCV into the pediatric NIP, pneumococcal disease caused by PCV serotype have declined among adults globally due to herd immunity (5–7). However, diseases caused by serotype 3 remain prevalent in many countries (5, 8, 9), likely due to the limited effectiveness of PCV against nasopharyngeal carriage of serotype 3 (22, 23). With the reduction of pneumococcal diseases caused by PCV-covered serotypes, pneumococcal disease caused by non-PCV serotypes have increased; emerging serotypes have varied by region or country. For example, serotype 8 has emerged as a common serotype in Europe, particularly in the UK, Spain, Italy (24–30), and South Africa (31). In England and Wales, serotype 8 accounted for more than 15% of IPD cases and pneumococcal pneumonia among individuals aged ≥15 years between 2016 and 2023 (25, 28, 29). Similarly, in South Africa, serotype 8 was the most common in IPD among adults aged ≥45 years during 2017–2019 (31). In contrast, serotype 4 has increased in the Western US and Canada among homeless and alcohol or drug-abusing individuals (32–34). However, the proportions of pneumococcal disease caused by serotype 8 and 4 remain low in Asian countries (35, 36), including Japan (16, 19). In our study, serotype 35B was the second most common serotype following serotype 3. This finding is consistent with previous research conducted in Japan. A study on the serotype distribution of IPD showed an increase in non-vaccine-covered serotypes, with a significant rise in serotype 35B (16). Another study investigating adult CAP in a single city in Japan reported serotype 35B as the most common, accounting for 12.0% of all pneumococcal CAP (37). Emerging serotypes following PCV introduction into the pediatric NIP vary across regions and countries. Therefore, domestic surveillance of pneumococcal disease is essential to monitor trends and modify vaccination policies effectively.

Although PPSV23 was introduced into NIP for adults aged ≥65 years in Japan, the proportion of the PPSV23 serotype identified in this study was 42.6% among adults aged ≥65 years. This may be attributed to low vaccine coverage (15, 16) or the limited effectiveness of PPSV23 in preventing pneumococcal pneumonia (38). PCV20 and PCV21 serotypes accounted for 42.8 and 72.3% of patients aged ≥65 years, respectively. With PCV15 and PCV20 now available for older adults, PCV21 currently under approval in Japan, and low pneumococcal vaccination coverage among adults, it is time to reconsider the routine pneumococcal vaccination strategy and explore ways to improve vaccination coverage. Furthermore, the introduction of PCV20 into the pediatric NIP in Japan in 2022 may influence the future serotype distribution associated with adult pneumococcal diseases. Therefore, ongoing surveillance and evaluation of pneumococcal disease trends are necessary.

Residents of nursing homes are at high risk of pneumococcal diseases due to having multiple underlying medical conditions and living in confined spaces (39, 40). When analyzing the pneumococcal serotype distribution separately for patients living in nursing homes and those living at home, the proportion of serotype 3 was higher in the former (25.9 versus 11.2%, respectively, p<0.01). Serotype 3 has been reported to cause more severe disease compared with other serotypes (41) and has been linked to clusters in nursing homes in Japan (42). From this perspective, pneumococcal vaccination that is expected to be effective against serotype 3 is particularly necessary for residents of nursing homes.

We detected 2.2% (13/583) of non-typable *S. pneumonia* isolates from adult pneumococcal pneumonia patients. While some isolations could not be serotyped, possibly due to technical issues, several may have been non-encapsulated *S. pneumoniae,* although this was not confirmed through genetic analysis. The polysaccharide capsule is a key pneumococcal virulence factor and the primary target for pneumococcal vaccines (43). Non-encapsulated *S. pneumoniae* has been reported to commonly cause otitis media and conjunctivitis or to be detected in carriage (44, 45). However, non-encapsulated *S. pneumoniae* can also cause IPD or pneumonia (16, 19, 46), posing a potential threat since currently used pneumococcal vaccines are not effective against non-encapsulated strains. Alternative vaccines that do not rely on targeting the polysaccharide capsule may therefore be required. It is essential to closely monitor trends in pneumococcal disease caused by non-encapsulated *S. pneumonia* to inform public health strategies effectively.

Our study had several limitations. Firstly, it included patients with pneumococcal pneumonia whose pneumococcal isolates were available for serotyping. Patients who were culture-negative but positive by polymerase chain reaction (PCR) or urinary assay were not included. It is possible that the use of alternative methods may yield different results (47). However, serotyping of culture-positive isolates using the Quellung reaction has been reported to be associated with acceptable levels of sensitivity and specificity (48), and we consider our study method reasonable for evaluating pneumococcal pneumonia serotypes. Secondly, due to the COVID-19 pandemic, criteria for sputum collection may have differed from those in the pre-pandemic period from an infection-control perspective, potentially resulting in the incomplete inclusion of pneumococcal pneumonia cases. However, when we compared the distribution of individual serotypes and proportion of vaccine-covered serotypes before (May 2019 to March 2020) and during (April 2020 to December 2022) the COVID-19 pandemic, no significant differences were observed, except for serotype 6C (Supplementary Table 3, Supplementary Figures 6 and 7). While different collection criteria may have influenced the number of pneumococcal isolates, it is unlikely that they affected the proportion of serotypes. Thirdly, although the proportion of serotype 3 was significantly higher in patients living in nursing homes compared with those living at home, the sample size for this subgroup was small.

## 5. Conclusion

In conclusion, serotype 3 and 35B were the most common pneumococcal serotypes identified in adult pneumococcal pneumonia patients in Japan between May 2019 and December 2022, with a higher proportion of serotype 3 observed among patients living in nursing homes compared with those living at home. Our findings revealed that serotypes included in PCV20 and PCV21 accounted for 42.8 and 72.3%, respectively, of patients aged ≥65 years, indicating that these new PCVs may offer additional protection against adult pneumococcal pneumonia.

## Acknowledgement

We thank Rina Shiramizu, Kyoko Uchibori, Yumi Araki, Yukie Kozone, Mao Hamasaki, and Eriko Yamada from the Institute of Tropical Medicine, Nagasaki University for their technical assistance. We also thank all the laboratory staff and the collaborators at the participating hospitals for their valuable contributions.

## Data availability

The data supporting the findings of this study are not available as consent for data sharing was not obtained.

## Author contributions

II and KM conceptualized and designed the study. HM, II, ES, NH, MS, IO, KN, HF, KO, TI, TY, SN, MT, YF, MT, YN, YH, AN, TS, AI, NS, YK, YT, TU, NA, AN, AY, TM, AU, MI, and KM collected the clinical data and bacterial strains. BGD and DON conducted the microbiological analysis. HM performed the data analysis and drafted the initial version of the manuscript. All authors provided critical review and revision of the text and approved the final version for publication.

## Abbreviations

CAP: community-acquired pneumonia
COVID-19: coronavirus disease 2019
IQR: interquartile range
IPD: invasive pneumococcal disease
NIP: national immunization programs
PCR: polymerase chain reaction
PCV: pneumococcal conjugate vaccines
PCV7: 7-valent pneumococcal conjugate vaccine
PCV13: 13-valent pneumococcal conjugate vaccine
PCV15: 15-valent pneumococcal conjugate vaccine
PCV20: 20-valent pneumococcal conjugate vaccine
PCV21: 21-valent pneumococcal conjugate vaccine
PPSV23: 23-valent pneumococcal polysaccharide vaccine.

