## Supplementary Material for "Serotype distribution among adults with community-acquired pneumococcal pneumonia in Japan between 2019 and 2022: A multicenter observational study"

### Supplementary Materials

**Table S1. Pneumococcal serotypes of adult pneumococcal pneumonia patients in Japan between 2019 and 2022, overall, by age group, and by place of residence**

| Pneumococcal serotype, no. (%) | All patients (n=583) | Age | | Place of residence | |
|---|---|---|---|---|---|
| | | Aged 18–64 years (n=132) | Aged ≥65 years (n=451) | Living at home (n=529) | Living in nursing homes (n=54) |
| 6A | 7 (1.2) | 0 | 7 (1.6) | 7 (1.3) | 0 |
| 4 | 1 (0.2) | 0 | 1 (0.2) | 1 (0.2) | 0 |
| 6B | 7 (1.2) | 1 (0.8) | 6 (1.3) | 4 (0.8) | 3 (5.6) |
| 9V | 0 | 0 | 0 | 0 | 0 |
| 14 | 1 (0.2) | 0 | 1 (0.2) | 0 | 1 (1.9) |
| 18C | 0 | 0 | 0 | 0 | 0 |
| 19F | 11 (1.9) | 2 (1.5) | 9 (2.0) | 11 (2.1) | 0 |
| 23F | 1 (0.2) | 0 | 1 (0.2) | 1 (0.2) | 0 |
| 1 | 0 | 0 | 0 | 0 | 0 |
| 3 | 73 (12.5) | 16 (12.1) | 57 (12.6) | 59 (11.2) | 14 (25.9) |
| 5 | 0 | 0 | 0 | 0 | 0 |
| 7F | 8 (1.4) | 2 (1.5) | 6 (1.3) | 8 (1.5) | 0 |
| 19A | 31 (5.3) | 10 (7.6) | 21 (4.7) | 28 (5.3) | 3 (5.6) |
| 22F | 16 (2.7) | 2 (1.5) | 14 (3.1) | 15 (2.8) | 1 (1.9) |
| 33F | 7 (1.2) | 3 (2.3) | 4 (0.9) | 7 (1.3) | 0 |
| 8 | 0 | 0 | 0 | 0 | 0 |
| 10A | 26 (4.5) | 6 (4.5) | 20 (4.4) | 25 (4.7) | 1 (1.9) |
| 11A | 39 (6.7) | 12 (9.1) | 27 (6.0) | 38 (7.2) | 1 (1.9) |
| 12F | 2 (0.3) | 0 | 2 (0.4) | 2 (0.4) | 0 |
| 15B | 25 (4.3) | 8 (6.1) | 17 (3.8) | 22 (4.2) | 3 (5.6) |
| 2 | 0 | 0 | 0 | 0 | 0 |
| 9N | 0 | 0 | 0 | 0 | 0 |
| 17F | 0 | 0 | 0 | 0 | 0 |
| 20 | 9 (1.5) | 3 (2.3) | 6 (1.3) | 9 (1.7) | 0 |
| 15A | 45 (7.7) | 8 (6.1) | 37 (8.2) | 39 (7.4) | 6 (11.1) |
| 15C | 13 (2.2) | 3 (2.3) | 10 (2.2) | 12 (2.3) | 1 (1.9) |
| 16F | 11 (1.9) | 3 (2.3) | 8 (1.8) | 10 (1.9) | 1 (1.9) |
| 23A | 37 (6.3) | 5 (3.8) | 32 (7.1) | 33 (6.2) | 4 (7.4) |
| 23B | 8 (1.4) | 0 | 8 (1.8) | 8 (1.5) | 0 |
| 24F | 10 (1.7) | 2 (1.5) | 8 (1.8) | 9 (1.7) | 1 (1.9) |

| | | | | | |
|---|---|---|---|---|---|
| 31 | 7 (1.2) | 1 (0.8) | 6 (1.3) | 7 (1.3) | 0 |
| 35B | 70 (12.0) | 17 (12.9) | 53 (11.8) | 67 (12.7) | 3 (5.6) |
| 13 | 1 (0.2) | 0 | 1 (0.2) | 1 (0.2) | 0 |
| 21 | 3 (0.5) | 1 (0.8) | 2 (0.4) | 3 (0.6) | 0 |
| 28 | 3 (0.5) | 0 | 3 (0.7) | 3 (0.6) | 0 |
| 29 | 8 (1.4) | 3 (2.3) | 5 (1.1) | 8 (1.5) | 0 |
| 34 | 31 (5.3) | 7 (5.3) | 24 (5.3) | 29 (5.5) | 2 (3.7) |
| 37 | 8 (1.4) | 1 (0.8) | 7 (1.6) | 7 (1.3) | 1 (1.9) |
| 38 | 3 (0.5) | 0 | 3 (0.7) | 3 (0.6) | 0 |
| 39 | 1 (0.2) | 0 | 1 (0.2) | 1 (0.2) | 0 |
| 40 | 1 (0.2) | 0 | 1 (0.2) | 1 (0.2) | 0 |
| 42 | 1 (0.2) | 1 (0.8) | 0 | 1 (0.2) | 0 |
| 24A | 2 (0.3) | 1 (0.8) | 1 (0.2) | 2 (0.4) | 0 |
| 24B | 2 (0.3) | 1 (0.8) | 1 (0.2) | 2 (0.4) | 0 |
| 33B | 1 (0.2) | 0 | 1 (0.2) | 1 (0.2) | 0 |
| 35A | 11 (1.9) | 3 (2.3) | 8 (1.8) | 10 (1.9) | 1 (1.9) |
| 35C | 3 (0.5) | 1 (0.8) | 2 (0.4) | 3 (0.6) | 0 |
| 35F | 4 (0.7) | 1 (0.8) | 3 (0.7) | 4 (0.8) | 0 |
| 6C | 15 (2.6) | 6 (4.5) | 9 (2.0) | 14 (2.6) | 1 (1.9) |
| 6D | 2 (0.3) | 1 (0.8) | 1 (0.2) | 2 (0.4) | 0 |
| 7C | 5 (0.9) | 0 | 5 (1.1) | 3 (0.6) | 2 (3.7) |
| Non-typable | 13 (2.2) | 1 (0.8) | 12 (2.7) | 9 (1.7) | 4 (7.4) |

**Table S2. Vaccine-covered pneumococcal serotypes of adult pneumococcal pneumonia patients in Japan between 2019 and 2022, overall, by age group, and by place of residence**

| Vaccine-covered serotypes, no. (%) | All patients (n=583) | Age | | Place of residence | |
|---|---|---|---|---|---|
| | | Aged 18–64 years (n=132) | Aged >65 years (n=451) | Living at home (n=529) | Living in nursing homes (n=54) |
| PCV13 | 140 (24.0) | 31 (23.5) | 109 (24.2) | 119 (22.5) | 21 (38.9) |
| PCV15 | 163 (28.0) | 36 (27.3) | 127 (28.2) | 141 (26.7) | 22 (40.7) |
| PCV20 | 255 (43.7) | 62 (47.0) | 193 (42.8) | 228 (43.1) | 27 (50.0) |
| PPSV23 | 257 (44.1) | 65 (49.2) | 192 (42.6) | 230 (43.5) | 27 (50.0) |
| PPSV23 non-PCV13 | 124 (21.3) | 34 (25.8) | 90 (20.0) | 118 (22.3) | 6 (11.1) |
| PCV21 | 419 (71.9) | 93 (70.5) | 326 (72.3) | 383 (72.4) | 36 (66.7) |
| PCV21 non-PCV20 | 210 (36.0) | 42 (31.8) | 168 (37.3) | 194 (36.7) | 16 (29.6) |
| PCV20 non-PCV21 | 46 (7.9) | 11 (8.3) | 35 (7.8) | 39 (7.4) | 7 (13.0) |
| Non PCV20/PCV21 | 118 (20.2) | 28 (21.2) | 90 (20.0) | 107 (20.2) | 11 (20.4) |

PCV13 serotype include serotype 4, 6 B, 9V, 14, 18C, 19F, 23F, 1, 5, 7F, 3, 6A, and 19A. PCV15 serotype include PCV13 serotype plus 22F and 33F. PCV20 serotype include PCV15 serotype plus 8, 10A, 11A, 12F, and 15 B. PPSV23 serotype include serotype 1, 2, 3, 4, 5, 6 B, 7F, 8, 9N, 9V, 10A, 11A, 12F, 14, 15 B, 17F, 18C, 19A, 19F, 20, 22F, 23F, and 33F. PCV23 non-PCV13 serotype include serotype 22F, 33F, 8, 10A, 11A, 12F, 15B, 2, 9N, 17F, and 20. PCV21 serotype include serotype 3, 6A, 7F, 19A, 22F, 33F, 8, 10A, 11A, 12F, 9N, 17F, 20, 15A, 15C, 16F, 23A, 23B, 24F, 31, and 35B. PCV21 non-PCV20 serotype include serotype 9N, 17F, 20, 15A, 15C, 16F, 23A, 23B, 24F, 31, and 35B. PCV20 non-PCV21 serotype include serotype 1, 4, 5, 6B, 9V, 14, 18C, 19F, 23F, and 15B. Non PCV21/PCV20 serotype, which are not included in PCV20 or PCV21, include serotype 2, 13, 21, 28, 29, 34, 37, 38, 39, 40, 42, 24A, 24B, 33B, 35A, 35C, 35F, 6C, 6D, 7C and non-typable.

Abbreviations: PCV13, 13-valent pneumococcal conjugate vaccine; PCV15, 15-valent pneumococcal conjugate vaccine; PCV20, 20-valent pneumococcal conjugate vaccine; PPSV23, 23-valent pneumococcal polysaccharide vaccine; PCV21, 21-valent pneumococcal conjugate vaccine.

**Table S3. Individual and vaccine-covered pneumococcal serotypes of adult pneumococcal pneumonia patients in Japan between 2019 and 2022, overall and by enrollment period**

| | All patients (n=583) | Enrollment period | |
|---|---|---|---|
| | | Before COVID-19 pandemic (n=275) | During COVID-19 pandemic (n=308) |
| **Individual serotypes, no. (%)** | | | |
| 6A | 7 (1.2) | 2 (0.7) | 5 (1.6) |
| 4 | 1 (0.2) | 1 (0.4) | 0 |
| 6B | 7 (1.2) | 3 (1.1) | 4 (1.3) |
| 9V | 0 | 0 | 0 |
| 14 | 1 (0.2) | 1 (0.4) | 0 |
| 18C | 0 | 0 | 0 |
| 19F | 11 (1.9) | 7 (2.5) | 4 (1.3) |
| 23F | 1 (0.2) | 1 (0.4) | 0 |
| 1 | 0 | 0 | 0 |
| 3 | 73 (12.5) | 34 (12.4) | 39 (12.7) |
| 5 | 0 | 0 | 0 |
| 7F | 8 (1.4) | 5 (1.8) | 3 (1.0) |
| 19A | 31 (5.3) | 17 (6.2) | 14 (4.5) |
| 22F | 16 (2.7) | 9 (3.3) | 7 (2.3) |
| 33F | 7 (1.2) | 3 (1.1) | 4 (1.3) |
| 8 | 0 | 0 | 0 |
| 10A | 26 (4.5) | 10 (3.6) | 16 (5.2) |
| 11A | 39 (6.7) | 21 (7.6) | 18 (5.8) |
| 12F | 2 (0.3) | 2 (0.7) | 0 |
| 15B | 25 (4.3) | 9 (3.3) | 16 (5.2) |
| 2 | 0 | 0 | 0 |
| 9N | 0 | 0 | 0 |
| 17F | 0 | 0 | 0 |
| 20 | 9 (1.5) | 6 (2.2) | 3 (1.0) |
| 15A | 45 (7.7) | 22 (8.0) | 23 (7.5) |
| 15C | 13 (2.2) | 6 (2.2) | 7 (2.3) |
| 16F | 11 (1.9) | 4 (1.5) | 7 (2.3) |
| 23A | 37 (6.3) | 13 (4.7) | 24 (7.8) |
| 23B | 8 (1.4) | 4 (1.5) | 4 (1.3) |
| 24F | 10 (1.7) | 6 (2.2) | 4 (1.3) |

| | | | |
|---|---|---|---|
| 31 | 7 (1.2) | 2 (0.7) | 5 (1.6) |
| 35B | 70 (12.0) | 28 (10.2) | 42 (13.6) |
| 13 | 1 (0.2) | 1 (0.4) | 0 |
| 21 | 3 (0.5) | 2 (0.7) | 1 (0.3) |
| 28 | 3 (0.5) | 3 (1.1) | 0 |
| 29 | 8 (1.4) | 0 | 8 (2.6) |
| 34 | 31 (5.3) | 14(5.1) | 17 (5.5) |
| 37 | 8 (1.4) | 4 (1.5) | 4 (1.3) |
| 38 | 3 (0.5) | 3 (1.1) | 0 |
| 39 | 1 (0.2) | 1 (0.4) | 0 |
| 40 | 1 (0.2) | 1 (0.4) | 0 |
| 42 | 1 (0.2) | 1 (0.4) | 0 |
| 24A | 2 (0.3) | 2 (0.7) | 0 |
| 24B | 2 (0.3) | 1 (0.4) | 1 (0.3) |
| 33B | 1 (0.2) | 1 (0.4) | 0 |
| 35A | 11 (1.9) | 7 (2.5) | 4 (1.3) |
| 35C | 3 (0.5) | 2 (0.7) | 1 (0.3) |
| 35F | 4 (0.7) | 1 (0.4) | 3 (1.0) |
| 6C | 15 (2.6) | 12 (4.4) | 3 (1.0) |
| 6D | 2 (0.3) | 0 | 2 (0.6) |
| 7C | 5 (0.9) | 2 (0.7) | 3 (1.0) |
| Non-typable | 13 (2.2) | 1 (0.4) | 12 (3.9) |
| **Vaccine-covered serotypes, no. (%)** | | | |
| PCV13 | 140 (24.0) | 71 (25.8) | 69 (22.4) |
| PCV15 | 163 (28.0) | 83 (30.2) | 80 (26.0) |
| PCV20 | 255 (43.7) | 125 (45.5) | 130 (42.2) |
| PPSV23 | 257 (44.1) | 129 (46.9) | 128 (41.6) |
| PCV21 | 419 (71.9) | 194 (70.5) | 225 (73.1) |

Before COVID-19 pandemic period defined as May 2019 to March 2020, while during COVID-19 pandemic period defined as April 2020 to December 2022. PCV13 serotype include serotype 4, 6 B, 9V, 14, 18C, 19F, 23F, 1, 5, 7F, 3, 6A, and 19A. PCV15 serotype include PCV13 serotype plus 22F and 33F. PCV20 serotype include PCV15 serotype plus 8, 10A, 11A, 12F, and 15 B. PPSV23 serotype include serotype 1, 2, 3, 4, 5, 6 B, 7F, 8, 9N, 9V, 10A, 11A, 12F, 14, 15 B, 17F, 18C, 19A, 19F, 20, 22F, 23F, and 33F. PCV21 serotype include serotype 3, 6A, 7F, 19A, 22F, 33F, 8, 10A, 11A, 12F, 9N, 17F, 20, 15A, 15C, 16F, 23A, 23B, 24F, 31, and 35B.

Abbreviations: COVID-19, coronavirus disease 2019; PCV13, 13-valent pneumococcal conjugate vaccine; PCV15, 15-valent pneumococcal conjugate vaccine; PCV20, 20-valent pneumococcal conjugate vaccine;

PPSV23, 23-valent pneumococcal polysaccharide vaccine; PCV21, 21-valent pneumococcal conjugate vaccine.

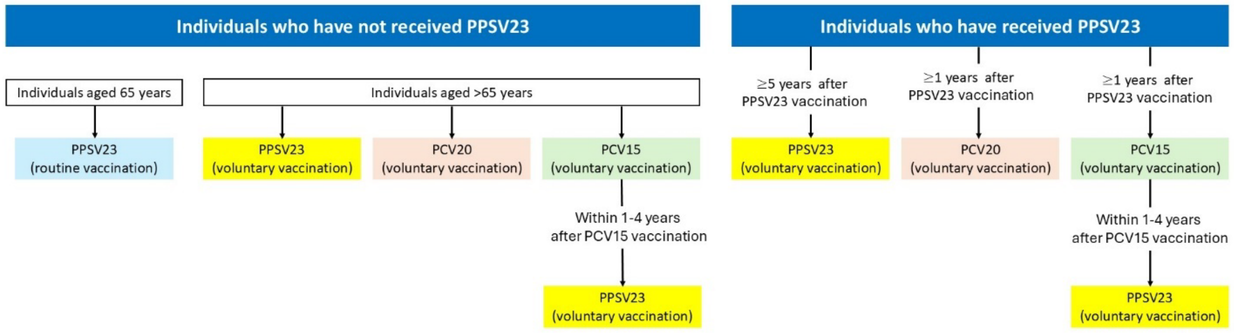

**Figure S1. Recommendations for pneumococcal vaccination of adults aged ≥65 years in Japan, issued by the Joint Committee of the Japanese Respiratory Society, Japanese Association for Infectious Diseases, and Japanese Society for Vaccinology.**

Abbreviations: PPSV23, 23-valent pneumococcal polysaccharide vaccine; PCV20, 20-valent pneumococcal conjugate vaccine; PCV15, 15-valent pneumococcal conjugate vaccine.

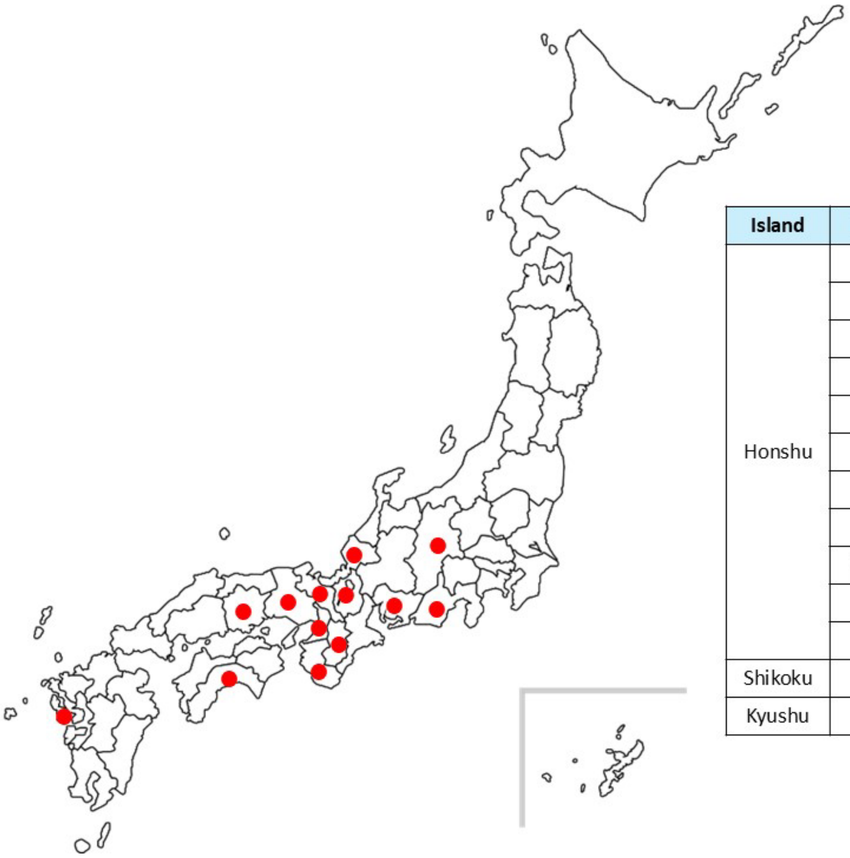

| Island | Prefecture | No. of hospitals |
|---|---|---|
| Honshu | Nagano | 1 |
| | Shizuoka | 1 |
| | Aichi | 1 |
| | Fukui | 1 |
| | Kyoto | 3 |
| | Shiga | 2 |
| | Osaka | 2 |
| | Nara | 2 |
| | Wakayama | 1 |
| | Hyogo | 4 |
| | Okayama | 1 |
| Shikoku | Kochi | 3 |
| Kyushu | Nagasaki | 2 |

**Figure S2. Prefectures in Japan with hospitals participating in this study.**

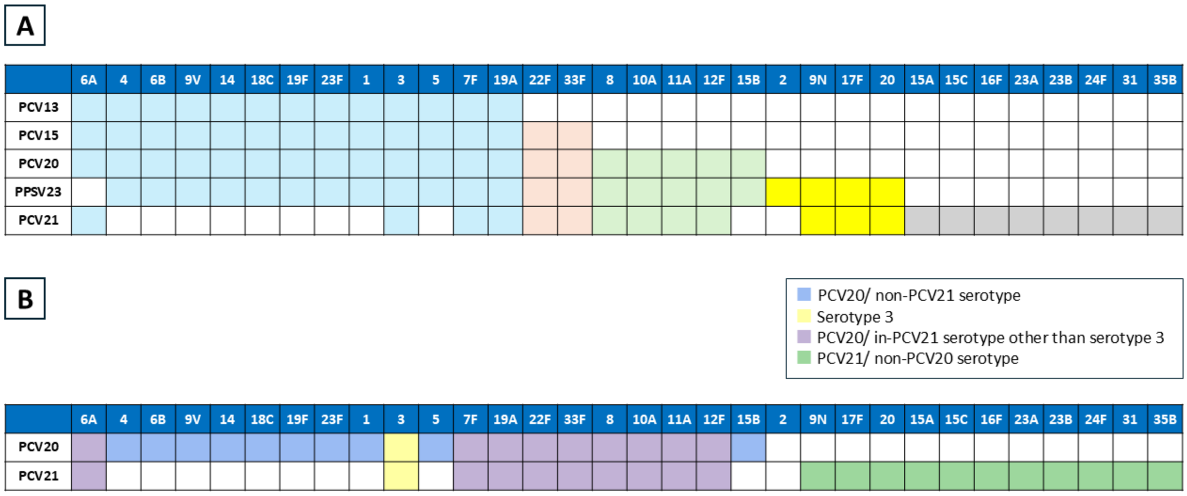

**Figure S3. Pneumococcal serotypes included in current and new pneumococcal vaccines.**

A. Pneumococcal serotypes included in PCV13, PCV15, PCV20, PPSV23, and PCV21. B. Pneumococcal serotypes included in PCV20 and PCV21 are shown to highlight the differences between PCV20 and PCV21. Abbreviations: PCV13, 13-valent pneumococcal conjugate vaccine; PCV15, 15-valent pneumococcal conjugate vaccine; PCV20, 20-valent pneumococcal conjugate vaccine; PPSV23, 23-valent pneumococcal polysaccharide vaccine; PCV21, 21-valent pneumococcal conjugate vaccine.

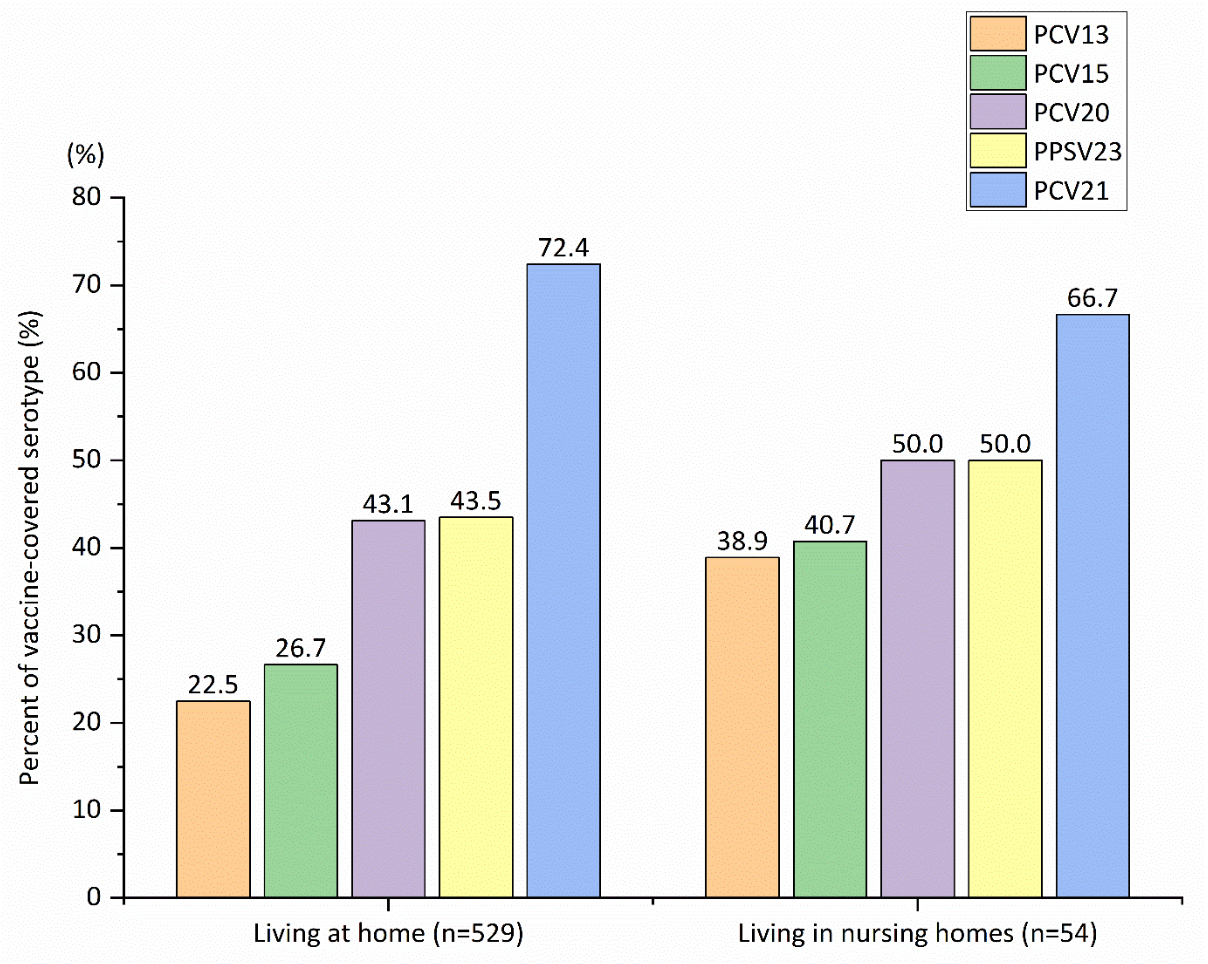

**Figure S4. Percentage of vaccine-covered serotypes among adult pneumococcal pneumonia patients in Japan between 2019 and 2022, categorized by place of residence.**

PCV13 serotype include serotype 4, 6 B, 9V, 14, 18C, 19F, 23F, 1, 5, 7F, 3, 6A, and 19A. PCV15 serotype include PCV13 serotype plus 22F and 33F. PCV20 serotype include PCV15 serotype plus 8, 10A, 11A, 12F, and 15 B. PPSV23 serotype include serotype 1, 2, 3, 4, 5, 6 B, 7F, 8, 9N, 9V, 10A, 11A, 12F, 14, 15 B, 17F, 18C, 19A, 19F, 20, 22F, 23F, and 33F. PCV21 serotype include serotype 3, 6A, 7F, 19A, 22F, 33F, 8, 10A, 11A, 12F, 9N, 17F, 20, 15A, 15C, 16F, 23A, 23B, 24F, 31, and 35B.

Abbreviations: PCV13, 13-valent pneumococcal conjugate vaccine; PCV15, 15-valent pneumococcal conjugate vaccine; PCV20, 20-valent pneumococcal conjugate vaccine; PPSV23, 23-valent pneumococcal polysaccharide vaccine; PCV21, 21-valent pneumococcal conjugate vaccine.

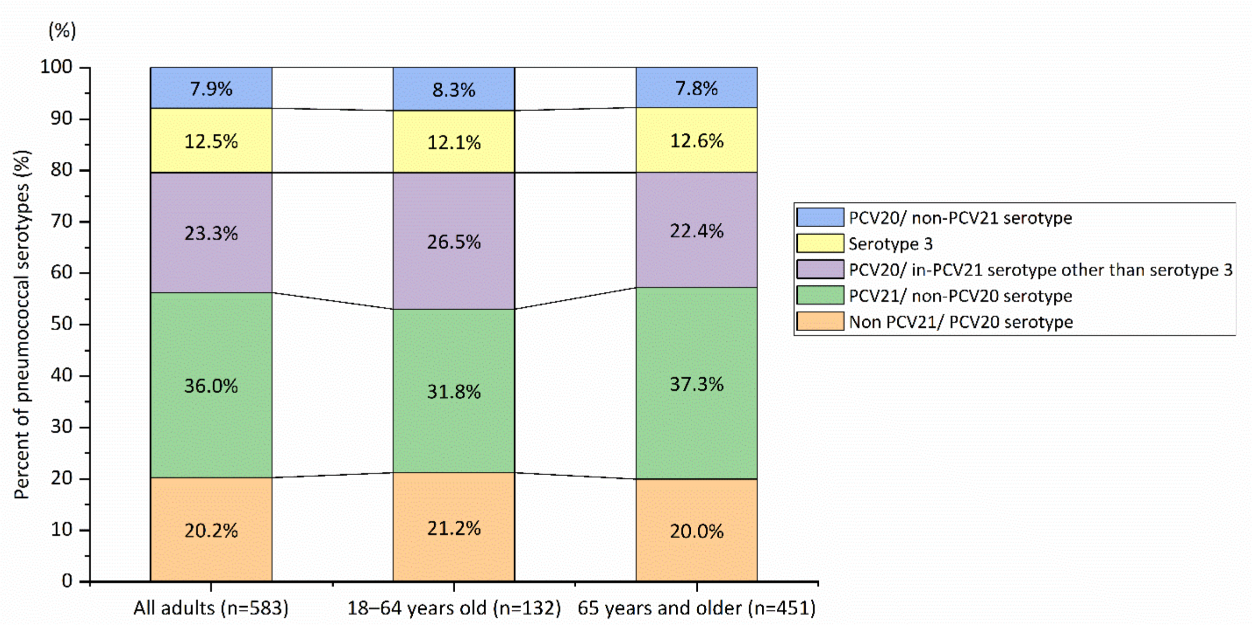

**Figure S5. Percentage of vaccine-covered serotypes among adult pneumococcal pneumonia patients in Japan between 2019 and 2022 overall, aged <65 years, and aged ≥65 years.**

PCV20/ non-PCV21 serotype include serotype 1, 4, 5, 6B, 9V, 14, 18C, 19F, 23F, and 15B. PCV20/ in-PCV21 serotype other than serotype 3 include serotype 6A, 7F, 19A, 22F, 33F, 8, 10A, 11A, 12F. PCV21/ non-PCV20 serotype include serotype 9N, 17F, 20, 15A, 15C, 16F, 23A, 23B, 24F, 31, and 35B. Non PCV21/PCV20 serotype, which are not included in PCV20 or PCV21, include serotype 2, 13, 21, 28, 29, 34, 37, 38, 39, 40, 42, 24A, 24B, 33B, 35A, 35C, 35F, 6C, 6D, 7C and non-typable.

Abbreviations: PCV20, 20-valent pneumococcal conjugate vaccine; PCV21, 21-valent pneumococcal conjugate vaccine.

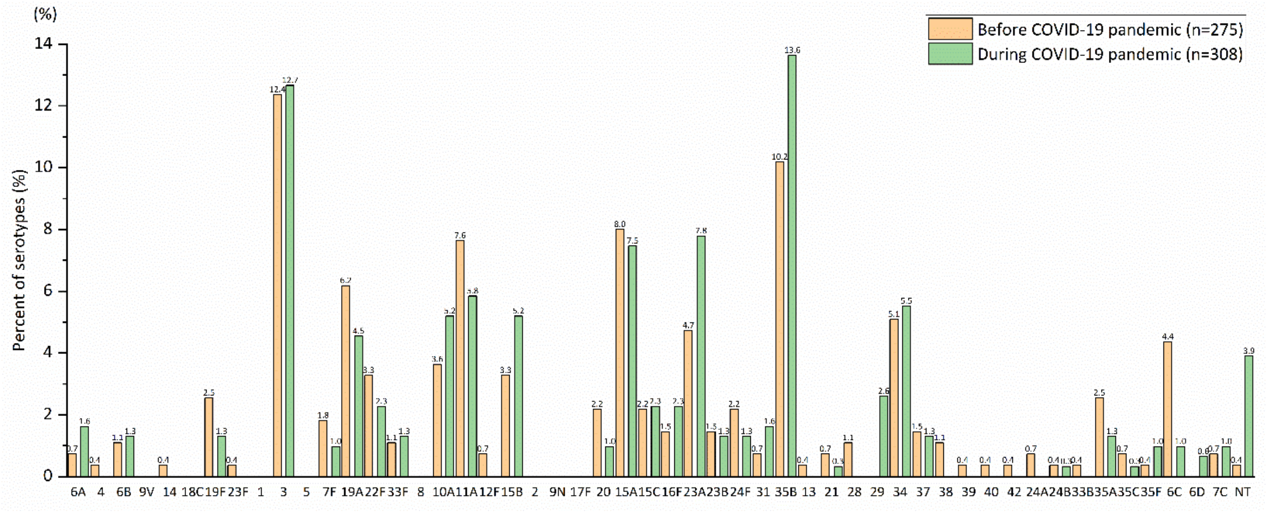

**Figure S6. Pneumococcal serotypes detected in adult pneumococcal pneumonia patients enrolled before (May 2019 to March 2020) or during (April 2020 to December 2022) the COVID-19 pandemic.**

The orange bar shows the percentage of pneumococcal serotypes detected in patients enrolled before the COVID-pandemic. The green bar shows that in patients enrolled during the COVID-19 pandemic.

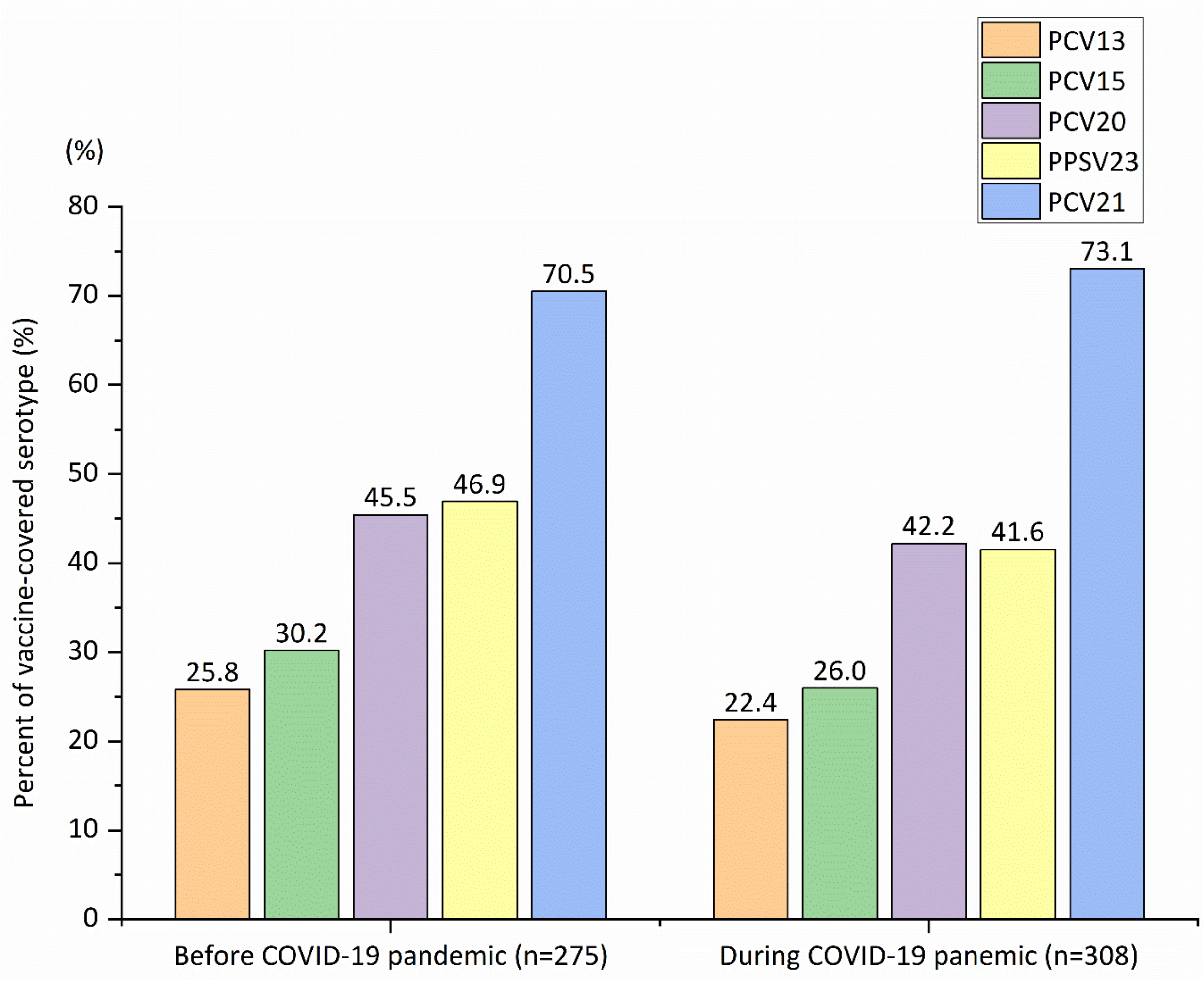

**Figure S7. Percentage of adult pneumococcal pneumonia patients with pneumococcal serotypes covered by PCV13, PCV15, PCV20, PPSV23, and PCV21 enrolled before (May 2019 to March 2020) or during (April 2020 to December 2022) the COVID-19 pandemic.**

PCV13 serotype include serotype 4, 6 B, 9V, 14, 18C, 19F, 23F, 1, 5, 7F, 3, 6A, and 19A. PCV15 serotype include PCV13 serotype plus 22F and 33F. PCV20 serotype include PCV15 serotype plus 8, 10A, 11A, 12F, and 15 B. PPSV23 serotype include serotype 1, 2, 3, 4, 5, 6 B, 7F, 8, 9N, 9V, 10A, 11A, 12F, 14, 15 B, 17F, 18C, 19A, 19F, 20, 22F, 23F, and 33F. PCV21 serotype include serotypes 3, 6A, 7F, 19A, 22F, 33F, 8, 10A, 11A, 12F, 9N, 17F, 20, 15A, 15C, 16F, 23A, 23B, 24F, 31, and 35B.

Abbreviations: PCV13, 13-valent pneumococcal conjugate vaccine; PCV15, 15-valent pneumococcal conjugate vaccine; PCV20, 20-valent pneumococcal conjugate vaccine; PPSV23, 23-valent pneumococcal polysaccharide vaccine; PCV21, 21-valent pneumococcal conjugate vaccine.
